# Sustained Effects of Low-to-Moderate Doses of Psilocybin on Brain Connectivity

**DOI:** 10.64898/2026.04.17.26351147

**Authors:** Chiranth Bhagavan, Orwa Dandash, Olivia Carter, Alexander Bryson, Richard A. Kanaan

## Abstract

**Background:** Psilocybin is a classic psychedelic that acutely alters brain functional connectivity. These changes are linked to therapeutic doses and subjective effects, with some evidence that changes persist beyond acute drug administration. However, the effects of lower doses on sustained connectivity changes remain unclear.

**Methods:** Ten healthy volunteers received three psilocybin doses (between 5 and 20 mg) in a randomized and blinded order, with at least one week between doses. Resting-state functional magnetic resonance imaging was completed at baseline and one week after a single dose. Functional connectivity changes were analyzed in relation to dose and altered conscious states at both the level of individual brain region connections (edges) and resting-state networks.

**Results:** Dose-dependent changes in 77 edges (76 increases, 1 decrease, of 1275 possible) were observed, but none survived multiple-comparison correction. At the network level, we observed one dose-dependent between-network increase (of 21 possible), and one dose-dependent within-network increase (of seven possible); the latter surviving correction. Alterations in conscious state were positively associated with widespread connectivity changes (dose-adjusted), with many network-level associations surviving correction. These directional patterns showed that lower doses and smaller conscious state alterations were linked to decreased connectivity, whereas higher doses and greater conscious state alterations were linked to increased connectivity.

**Conclusions:** Dose level and acute subjective effects were positively associated with multiple functional connectivity changes one week after a low-to-moderate psilocybin dose. Further research is warranted to characterize these sustained effects and their therapeutic relevance to inform studies adopting similar dosing regimens in clinical cohorts.

**Trial Registration:** Australian New Zealand Clinical Trials Registry: ACTRN12621000560897

Date registered: 12 May 2021

URL: https://www.anzctr.org.au/Trial/Registration/TrialReview.aspx?id=381526&isReview=true

## Introduction

Psilocybin is a classic psychedelic associated with widespread effects on the brain that are thought to arise primarily through partial agonism of the serotonin-2A receptor (Nichols, 2016). These effects range from changes in structural and functional brain connectivity (Kringelbach et al., 2020; Siegel et al., 2024) to acutely altered conscious states (Yaden et al., 2021) and enduring changes in beliefs and behaviors (Studerus et al., 2011). Supported by encouraging treatment outcomes in neuropsychiatric disorders (Andersen et al., 2021), contemporary models for these diverse effects propose that psilocybin increases neuroplasticity and sensitivity to sensory input (Carhart-Harris and Friston, 2019; van Elk and Yaden, 2022).

Resting-state functional magnetic resonance imaging (fMRI) studies provide some support towards these proposed therapeutic effects. A recent systematic review reported consistent findings of reduced functional connectivity within the default mode network and increased connectivity between resting-state networks (Berkovitch et al., 2025). These findings have often been associated with plasma drug levels and the subjective intensity of drug effects, implicating both as possible correlates of connectivity changes (Carhart-Harris et al., 2012; Lebedev et al., 2016; Madsen et al., 2021; Preller et al., 2018; Siegel et al., 2024; Tolle et al., 2024). However, most studies focussed on acute connectivity changes within 24 hours of drug administration, whereas current frameworks for psychedelic treatment emphasise active therapy in the weeks following dosing, based on hypothesised ‘persisting’ neuroplastic effects and capacity for change (Barber and Aaronson, 2022; Passie et al., 2022; Watts and Luoma, 2020; Weiss et al., 2025).

Only six studies (from five datasets) investigated persisting effects of psilocybin on resting-state functional connectivity (Barrett et al., 2020; Daws et al., 2022; Deco et al., 2024; Doss et al., 2021; McCulloch et al., 2022a; Siegel et al., 2024), ranging from one week to twelve months post-drug administration, and all involving moderate-to-high doses. Compared to the evidence of psilocybin’s acute effects on brain function, these studies provide less evidence for sustained changes within the default mode network, and inconsistent evidence of cross-network connectivity changes and relevant therapeutic correlates.

Crucially, sustained connectivity changes following lower doses have not been explored, despite growing clinical interest in this treatment approach. Moreover, given the potential for dose-dependent adverse effects (Romeo et al., 2024), lower doses may offer improved tolerability, potentially extending accessibility to individuals who have typically been excluded from high-dose clinical studies. Studies of low-to-moderate psilocybin doses have shown encouraging outcomes in cluster headache (Schindler et al., 2022), migraine (Schindler et al., 2021), and post-treatment Lyme disease (Garcia-Romeu et al., 2026), with further studies underway to investigate fibromyalgia and motor functional neurological disorder (FND) (Bhagavan et al., 2025, 2026). Further research is required to better understand persisting changes in functional connectivity following low-to-moderate doses to inform these expanding treatment applications.

In this resting-state fMRI study, we investigate changes in functional connectivity in healthy volunteers one week after the first of three low-to-moderate psilocybin doses administered in a randomised order. Given the range of doses, we focused on the relationship between drug dose, experiential intensity, and any persisting effects of psilocybin on brain connectivity. Overall group-level analyses, without modeling these covariates, are also provided to explore whether connectivity changes persist across varying dose levels and subjective effects. Analyses were conducted between individual regions of interest (ROIs) to identify specific brain regions driving connectivity changes, and across resting-state networks to characterize effects on global network architecture, including distributed effects which may be more likely to aggregate across participants at the network level (Chopra et al., 2024; Segal et al., 2023). We hypothesised that both dose and the degree of alteration in subjective experience would be positively associated with connectivity increases across several brain regions and networks.

## Methods and Materials

This fMRI investigation was conducted as part of a triple-blind, randomized, dose-finding pilot study assessing the impact of low-to-moderate doses of psilocybin on motor function in healthy individuals, in part to inform a subsequent, Phase 1 study of psilocybin-assisted physiotherapy in motor FND (Bhagavan et al., 2025). Ethical approval was granted by the Austin Health Human Research Ethics Committee (HREC/57390/Austin-2020). The trial was registered on the Australian New Zealand Clinical Trials Registry, ACTRN12621000560897, and the study protocol was published, outlining full details of participant eligibility and the study design (Bhagavan et al., 2024).

### Participants

Eligible participants were healthy adults aged 18 to 65, excluding those with medical or psychiatric conditions or medications that might pose risks following psilocybin administration.

### Study Sequence

Twelve participants took part in the pilot study and were divided into two blocks by order of enrolment:

- The first six participants were assigned to take oral psilocybin 5, 10, and 15 mg.
- The next six participants were assigned to take oral psilocybin 10, 15, and 20 mg.

The participants’ order of dosing was randomized with one week scheduled between doses, and they remained blinded to the order of dosing throughout enrolment. At the end of each psilocybin dosing session, participants were asked to guess the dose received, and altered conscious states were assessed by administering the 5-Dimensional Altered States of Consciousness (de Deus Pontual et al., 2023) and Ego-Dissolution Inventory (Nour et al., 2016), with mean scores computed for each measure.

fMRI was acquired at baseline on the morning of their first dose prior to drug administration, and follow-up imaging was scheduled one week later, just before their second dose (Figure 1). Only one follow-up fMRI one week after the initial dose was scheduled due to budget constraints and to minimize additional study visits and participant burden. Each participant, therefore, contributed observations from a single dosing session, enabling group-level analyses across varying dose levels and phenomenological intensities due to the randomized dose order, while within-subject association analyses were not feasible.

**Figure 1:**
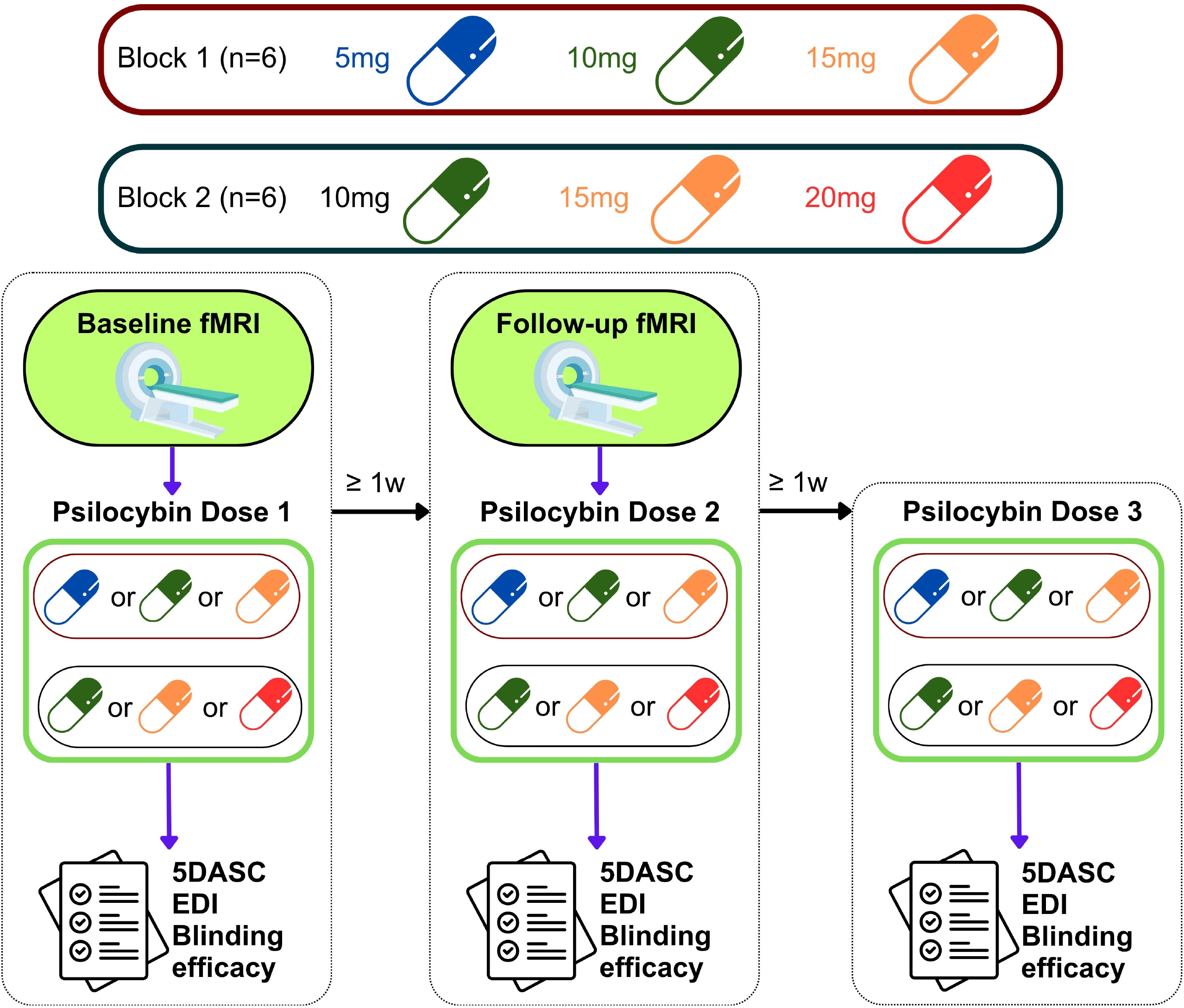
Study schedule. fMRI = functional magnetic resonance imaging 1w = 1 week 5DASC = 5-Dimensional Altered States of Consciousness EDI = Ego-Dissolution Inventory

### Neuroimaging acquisition, quality control, and preprocessing

Images were obtained on a Siemens 3 Tesla wide-bore Vida magnetic resonance imaging (MRI) system fitted with a 64-channel BioMatrix head coil. Participants first underwent a localizer scan to allow the scanner to determine brain position within the magnet. This was followed by a high-resolution T1-weighted structural scan for functional time series co-registration. Participants then completed a 15-minute, T2*-weighted resting-state fMRI scan with their eyes closed and without music.

Each participant’s data was visually inspected for obvious artefacts or lesions. Image preprocessing was performed in Statistical Parametric Mapping software (SPM12), running in Matlab (MathWorks, Natick, MA, USA). This involved head-motion correction, co-registration with the T1 structural weighted image, and then normalization to a common standard space defined by the Montreal Neurological Institute (MNI) 152 template (Mazziotta et al., 2001). The anatomical images then underwent skull stripping and segmentation of tissue types. Signal from white matter, cerebrospinal fluid, and the global brain signal, in addition to the 6 head-motion parameters and their first derivatives, were modelled as nuisance variables and removed from the analysis. Regular visual quality control to ensure correct co-registration and normalization was conducted. The functional data underwent linear detrending, bandpass filtering at 0.008-0.08 Hz, and spatial smoothing with a Gaussian kernel (full width at half maximum) of 8 mm.

Detailed information regarding neuroimaging acquisition, quality control, and preprocessing is provided in the Supplementary Information.

### First-level modeling

The pre-processed and denoised datasets were parcellated into 51 ROIs, centered on centroid coordinates of split components derived from the Yeo functional atlas (Wang et al., 2019; Yeo et al., 2011), providing representative sampling of each of the seven canonical resting-state networks. The ROIs were built using MaRSBaR (http://marsbar.sourceforge.net), defined by a 6mm sphere about their MNI coordinates.

The first eigenvariables were extracted for each ROI’s timeseries at each of the 300 volumes acquired to produce a matrix of 51 x 300 timeseries for each acquisition. Given the small sample size, Spearman rank correlations were used to estimate functional connectivity, as these require fewer assumptions regarding the distribution of blood oxygen level-dependent timeseries data, and to reduce sensitivity to outliers that may exert undue influence in smaller datasets. Spearman rank correlations were computed to determine the connectivity strength for each ROI pair, or edge, providing a 51 x 51 matrix. To stabilize variance, Spearman correlation coefficients were clipped (−0.999999 to 0.999999) and Fisher-transformed to z-scores. Duplicate edges were excluded so that the functional connectivity change for each edge was only counted once, resulting in 1275 possible edges. This procedure was repeated for each participant at each acquisition timepoint (baseline and follow-up). Within-participant functional connectivity change was computed by subtracting baseline from follow-up z-scores. This provided one value per participant, preserving dependence within individual timeseries and paired-fMRI acquisitions, enabling exchangeability and subsequent permutation-based testing.

To explore effects on global network architecture, connectivity within and between the seven resting-state networks was assessed, hereafter referred to as ‘network-level’ analyses. For within-network functional connectivity change, the average change in connectivity of all edges comprising ROIs belonging to the same network was calculated. Between-network functional connectivity change was calculated by averaging the connectivity change for each edge connecting ROIs between two networks. This procedure was repeated for each participant, resulting in average within- and between-network functional connectivity changes per participant.

### Second-level modeling

Permutation analyses were performed to identify functional connectivity changes one week post-psilocybin administration compared to baseline. This statistical approach is well established and offers advantages in neuroimaging analyses, given the minimal assumptions regarding data distribution in small sample sizes and more reliable control of false positives (Dellert et al., 2025; Ely et al., 2019; Orr et al., 2020; Quent et al., 2025; Williams et al., 2025; Winkler et al., 2014; Wong et al., 2024).

#### Dose-response

To model dose-dependent effects, ordinary least squares regression was used to estimate a slope reflecting functional connectivity change with every 5 mg increase in dose. The observed test statistic was derived by the slope divided by its standard error. Dose values were randomly shuffled via 100,000 permutations across the participants, with a regression model and test statistic recomputed for each permutation. This provided an empirical null distribution corresponding to the hypothesis that there was no association between dose and functional connectivity change. Two-sided, uncorrected permutation *p*-values (*p*_unc_) were calculated as the proportion of permuted test statistics greater than or equal to the value of the actual, observed statistic, via:

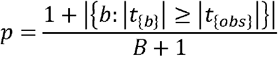

where *p* is *p*_unc_, *t*_{b}_ is the *t*-statistic computed from permutation *b, t*_{obs}_ is the *t*-statistic computed from the observed data, and *B* is the number of permutations.

#### Associations with altered conscious states

To examine associations between altered conscious states and functional connectivity change beyond the effects of dose, 5-Dimensional Altered States of Consciousness and Ego-Dissolution Inventory scores and connectivity changes were residualized with respect to the mean-centered psilocybin dose. Associations between these residuals were then estimated using ordinary least squares regression, with the test statistic derived by dividing the regression slope by its standard error. Behavioral residuals were randomly shuffled via 100,000 permutations across the participants, with a regression model and test statistic recomputed for each permutation. This provided an empirical null distribution corresponding to the hypothesis that there was no association between altered conscious state and functional connectivity change after adjusting for dose. Two-sided, uncorrected permutation *p*-values (*p*_unc_) were calculated, as described above.

A sensitivity test was performed for the association between network-level functional connectivity and altered conscious states without adjusting for dose to assess the general consistency and suitability of the dose-adjusted analysis.

#### Group-level change

A sign-flip permutation test on within-participant functional connectivity changes was performed by establishing all possible assignments (2^n^) across all participants, corresponding to the null hypothesis that the distribution of connectivity change was symmetrical around zero. For each test, the observed group-level statistic was derived by the group-level mean functional connectivity change divided by its standard error, providing a one-sample *t*-statistic. For each permutation, participant-wise functional connectivity change scores were multiplied by ±1 before recomputing the statistic. Two-sided, uncorrected permutation *p*-values (*p*_unc_) were calculated, as described above.

For each of the above analyses, the permutation-based procedure was applied to edgewise, within-network, and between-network connectivity changes. Correction for multiple comparisons was applied to unadjusted *p*-values within each analysis by controlling for the false discovery rate (FDR) at 5% using the Benjamini-Hochberg procedure to provide FDR-adjusted *p*-values (*p*_FDR_) (Benjamini and Hochberg, 1995). Given the small sample size and exploratory nature of this study, results surviving an uncorrected threshold of *p* < 0.05 are reported and are denoted as ‘*p*_unc_ < 0.05’ for transparency. Where applicable, findings meeting *p*_FDR_ < 0.05 are indicated as ‘surviving correction’.

Details of the study design, including timepoints for fMRI acquisitions, dosing, and behavioral measures, are provided in Figure 1.

## Results

### Participants

Thirteen participants were enrolled, of whom three were excluded from the imaging analysis. One was withdrawn due to a study protocol breach, did not complete either scan, and was replaced; one did not complete either scan; and one completed their follow-up scan two months post-dose, substantially exceeding the scheduled one-week post-dose timing (Figure S1). The final sample for this study was therefore 10 (mean age [range] = 30.1 [22-39] years; mean height [range] = 170.8 [160-188] cm; mean weight [range] = 68.5 [51-94] kg; 5 (50%) females, 5 (50%) males) (Table S2A). For three participants, substantial time elapsed between their first and second dose, when the follow-up scan was planned, so their follow-up scan was rescheduled to one week after their second dose (just prior to their third dose) to maintain consistency in the one-week post-dose timing of the follow-up scan. Follow-up scans were completed seven days post-dose for nine participants and six days post-dose for the remaining participant. Follow-up scans were acquired following a 5 mg dose in one participant, 10 mg in three participants, 15 mg in two participants, and 20 mg in four participants (mean dose ± standard deviation = 14.5 ± 5.2 mg) (Table S2B). Four participants (40%) correctly guessed the dose preceding their follow-up scan, which was only slightly above chance (33.3%).

### Dose-dependent effects

77 of the possible 1275 edges demonstrated dose-dependent functional connectivity change (*p*_unc_ < 0.05) but did not survive correction for multiple comparisons. 76 of these edges showed dose-dependent increases in connectivity between ROIs both within and between several resting-state networks. The remaining edge demonstrated a dose-dependent decrease in connectivity between two ROIs within the frontoparietal control network (Figure 2A).

**Figure 2:**
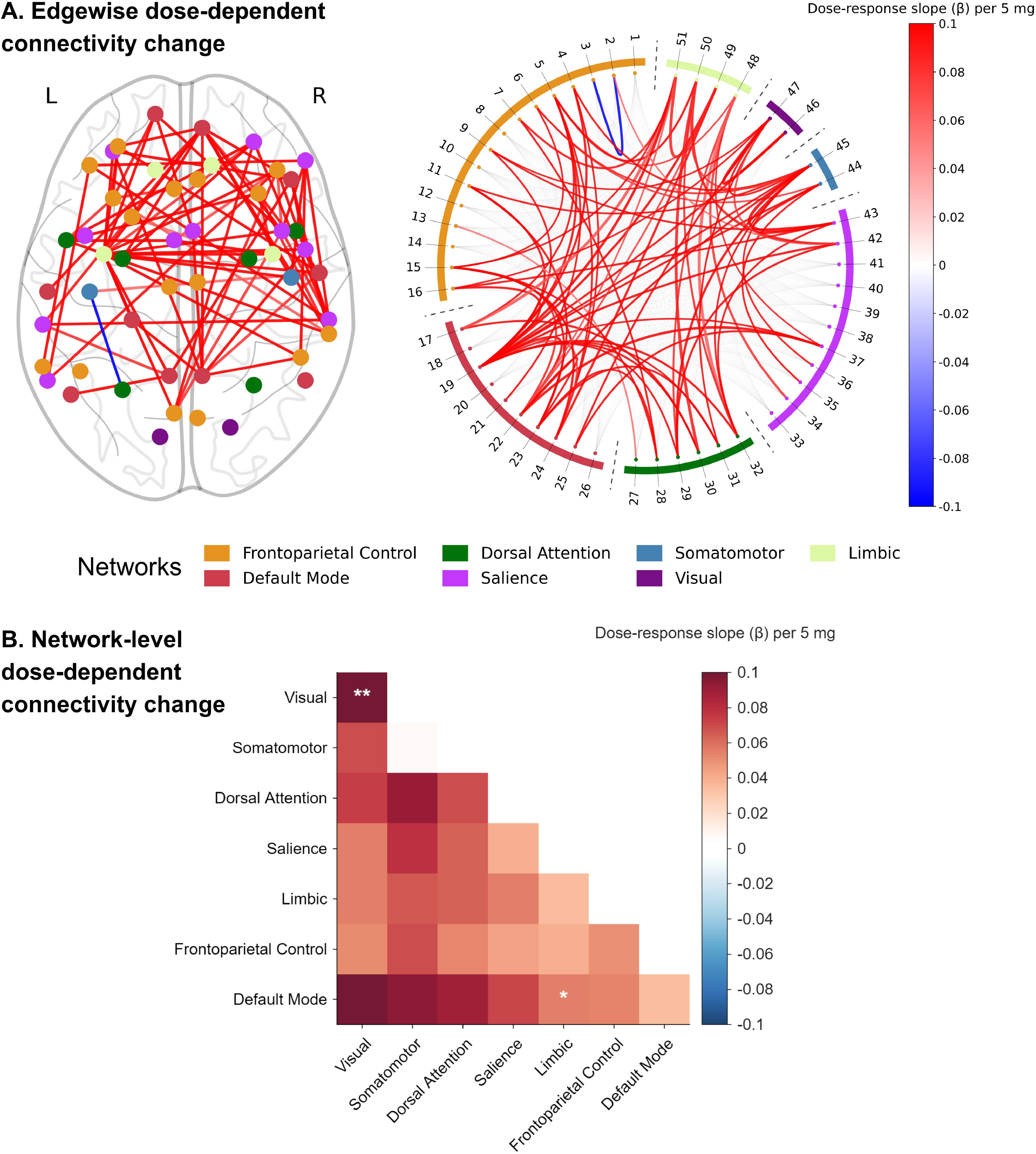
Dose-dependent functional connectivity change. A: Edgewise functional connectivity changes at follow-up compared to baseline as a function of dose. Results are viewed at *p*_*unc*_ < 0.05. Region of interest (ROI) color is represented by its corresponding network. Edge color is represented by the dose-response slope (β) per 5 mg (see color bar on the right). Relative edge width is represented by the t-statistic. See Table S1 for ROI labels. B: Network-level functional connectivity changes at follow-up compared to baseline as a function of dose. Square color is represented by the dose-response slope (β) per 5 mg (see color bar on the right). The diagonal squares represent within-network change, while the off-diagonal squares represent between-network change. *\*p*_*unc*_ < 0.05; *\*\*p*_*FDR*_ < 0.05.

Aggregated at the network level (*p*_unc_ < 0.05), a dose-dependent increase in connectivity was observed between the visual and somatomotor networks (of a possible 21 between-network connections) and a dose-dependent increase was observed within the visual network (of a possible seven within-network connections), the latter surviving correction for multiple comparisons (Figure 2B).

### Associations with altered conscious states

Functional connectivity change for 364 of the possible 1275 edges showed associations with the 5-Dimensional Altered States of Consciousness, and 217 edges showed associations with the Ego-Dissolution Inventory, after adjusting for dose (*p*_unc_ < 0.05). Three of the 364 associations with the 5-Dimensional Altered States of Consciousness that survived *p*_unc_ < 0.05 were negative, including one edge spanning ROIs within the default mode network, and the remaining two edges each spanning one ROI from the default mode network and one ROI from the limbic network. In contrast, all remaining associations across both behavioural measures that survived *p*_unc_ < 0.05 were positive, spanning ROI pairs both within and between several resting-state networks (Figure S2). Although none of these edgewise associations survived correction for multiple comparisons, the consistent, positive direction of effects warrants noting in the context of this small sample pilot study.

Aggregated at the network level, similarly widespread positive associations were observed between altered conscious states and functional connectivity change, after adjusting for dose. The 5-Dimensional Altered States of Consciousness showed positive associations with functional connectivity change for 18 between-network pairs (*p*_unc_ < 0.05), with each of the seven resting-state networks represented in at least one pair, and 16 of which survived correction for multiple comparisons. Connectivity changes for three within-network pairs, comprising the visual, dorsal attention, and salience networks, also demonstrated positive associations, all of which survived correction.

For the Ego-Dissolution Inventory, network-level positive associations with connectivity change were observed for seven between-network pairs (*p*_unc_ < 0.05), involving the visual, somatomotor, salience, dorsal attention, and frontoparietal control networks in at least one pair; however, none of these pairs survived correction. Positive associations were also observed for five within-network pairs, comprising the visual, somatomotor, dorsal attention, frontoparietal control, and default mode networks, all of which survived correction. Across both measures of altered conscious state, no negative associations were observed for either within- or between-network connectivity changes (Figure 3B).

**Figure 3:**
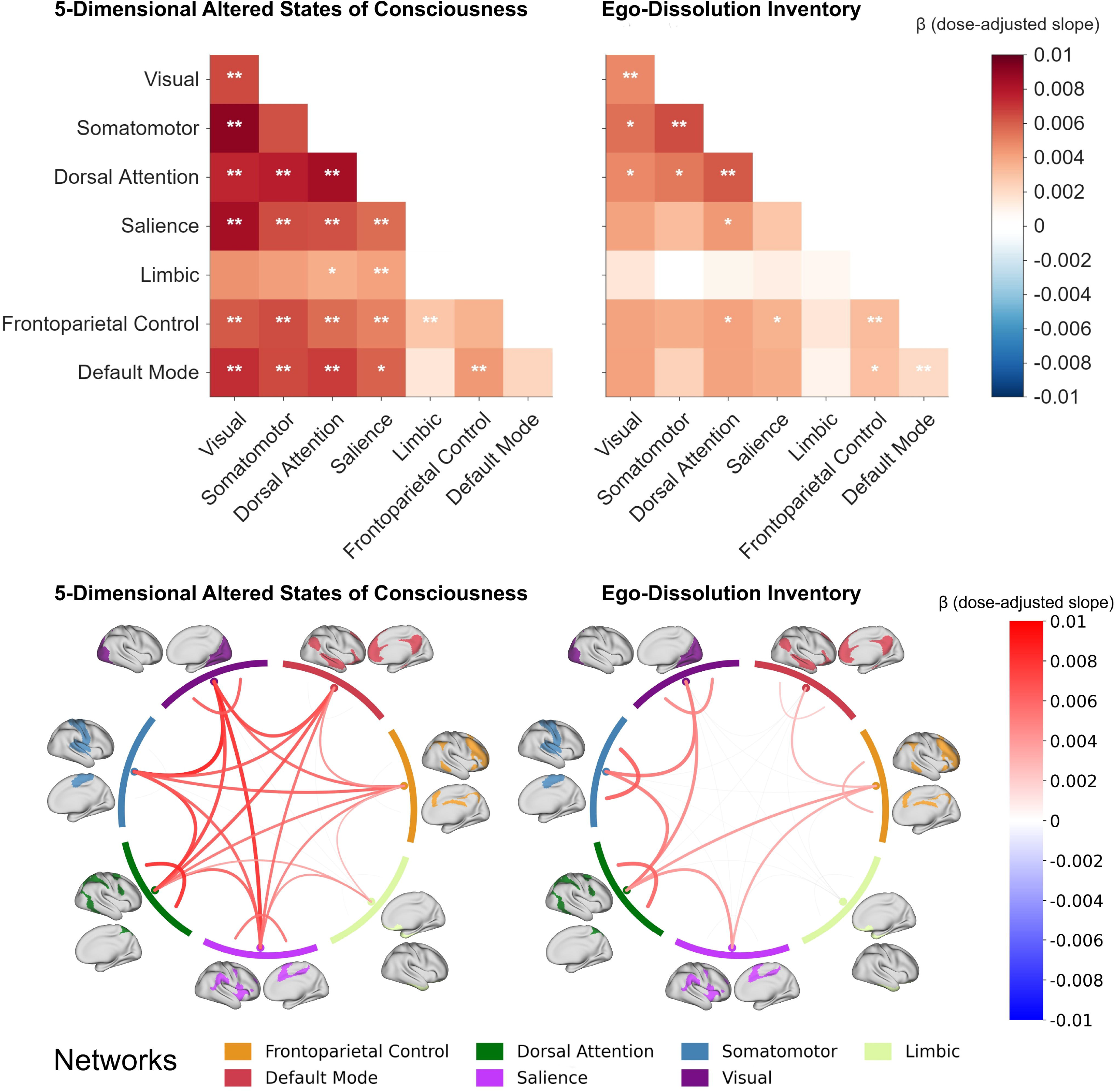
Network-level functional connectivity changes as a function of altered conscious states (dose-adjusted) Functional connectivity changes at follow-up compared to baseline as a function of the mean 5-Dimensional Altered States of Consciousness (left) and mean Ego-Dissolution Inventory (right) (dose-adjusted): • Heatmap: Square color is represented by the slope (β) (see color bar on the right). The diagonal squares represent within-network change, while the off-diagonal squares represent between-network change. *\*p*_*unc*_ < 0.05; *\*\*p*_*FDR*_ < 0.05. • Chord chart: Edge color is represented by slope (see color bar on right). Relative edge width is represented by the t-statistic. Results are viewed at *p*_*unc*_ < 0.05.

The sensitivity analysis of unadjusted network-level associations between functional connectivity change and altered conscious states was broadly consistent with these dose-adjusted findings (Figure S3). This showed positive associations between several within- and between-network functional connectivity changes and both the 5-Dimensional Altered States of Consciousness and Ego-Dissolution Inventory, reinforcing the suitability of the dose-adjusted findings for exploring these associations beyond the dose-effect.

### Directional patterns

Synthesising these associations across dose and altered conscious states, for all network-level associations surviving *p*_unc_ < 0.05, positive correlations were consistently observed. This showed that lower doses and subjective intensity were often accompanied by several decreases in functional connectivity at follow-up compared to baseline, while higher doses and higher alterations in conscious states were often accompanied by increases in functional connectivity. All network-level associations with dose and altered conscious states (dose-adjusted) are provided in the Supplementary Information (Figures S4-6). Figure 4 provides an abridged visualisation of the two most statistically significant associations between network-level functional connectivity change and (A) dose and (B-C) altered conscious states (dose-adjusted) to illustrate this directional pattern.

**Figure 4:**
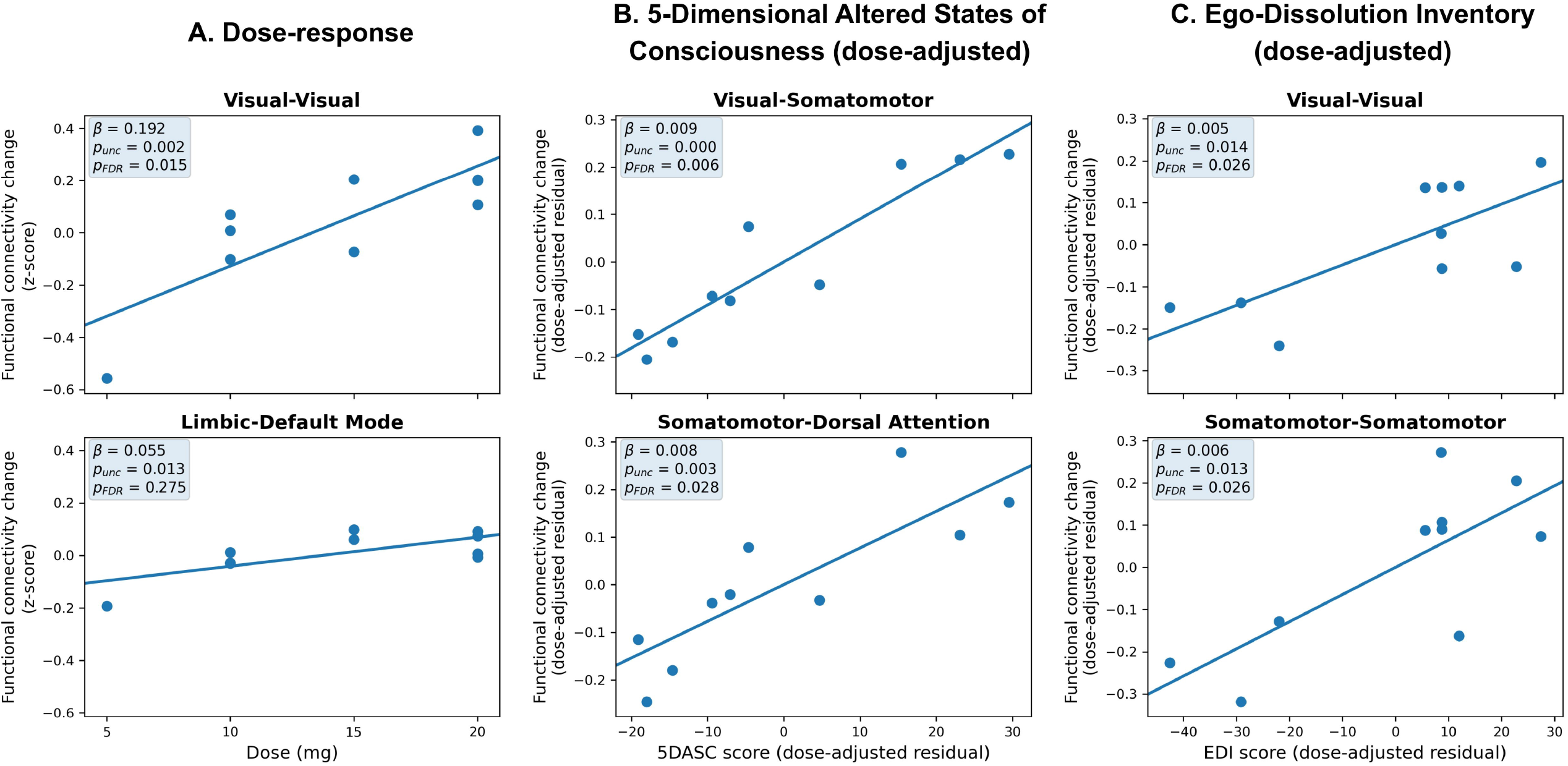
Associations between dose, altered conscious states (dose-adjusted), and network-level functional connectivity change. A: Two most statistically significant network-level functional connectivity changes at follow-up compared to baseline as a function of dose (per 5 mg). B: Two most statistically significant network-level functional connectivity changes at follow-up compared to baseline as a function of mean 5-Dimensional Altered States of Consciousness (5DASC) score (dose-adjusted). C: Two most statistically significant network-level functional connectivity changes at follow-up compared to baseline as a function of mean Ego-Dissolution Inventory (EDI) score (dose-adjusted).

### Group-level functional connectivity change

Pooled across all participants, ten of the possible 1275 edges demonstrated functional connectivity change (*p*_unc_ < 0.05) but did not survive correction for multiple comparisons. Two of these edges represented decreased connectivity: one edge corresponding to two ROIs within the default mode network, and the other between two ROIs from the salience and limbic networks, respectively. Of the eight edges that showed increased connectivity, two edges represented within-network connectivity increases for ROIs within the dorsal attention and frontoparietal control networks, respectively, while the remaining six edges represented increased between-network connections (Figure S7A).

Aggregated at the network-level, no group-level within- or between-network connectivity changes were significant at a statistical threshold of *p*_unc_ < 0.05 (Figure S7B).

## Discussion

This resting-state fMRI study found that a subset of persisting functional connectivity changes one week after a low-to-moderate psilocybin dose were positively associated with dose and acutely altered conscious states. These findings highlight the potential relevance of dosing and experiential factors in shaping the desired direction of sustained connectivity change. However, the study is limited by a small sample size and low statistical power, with only a portion of observed associations surviving correction for multiple comparisons.

These positive associations share some overlap with the existing literature exploring the effects of higher psychedelic doses on acute changes in brain connectivity. Plasma psilocin level (the active metabolite of psilocybin) and subjective drug intensity have been positively correlated with widespread, acute changes in between-network connectivity, particularly between the default mode and dorsal attention networks and other cortical networks (Madsen et al., 2021). A study of lysergic acid diethylamide (another psychedelic agent) in healthy individuals found positive associations between the 5-Dimensional Altered Conscious States and acute changes in somatomotor network connectivity with the rest of the brain (Preller et al., 2018), while another study of psilocybin in healthy individuals demonstrated positive associations with the Mystical Experiences Questionnaire and acute connectivity changes involving the association cortex, whereas primary sensorimotor regions were relatively spared (Siegel et al., 2024). Many of these findings align with our observed correlations with persisting connectivity increases between these cortical networks. Moreover, our study expands on this literature by including lower doses, thereby reinforcing these associations across varying levels of pharmacological and phenomenological intensity.

Analysis of these associations is lacking in previous studies of sustained connectivity changes. One study detected no significant correlations between acutely altered conscious states and connectivity change within the executive control network at one week or three months post-dose, and correlations with other regions or networks were not analyzed (McCulloch et al., 2022a). No other fMRI study of sustained changes in resting-state connectivity following psilocybin administration that we identified reported correlations with dose or acutely altered conscious states. This study’s findings, therefore, provide novel evidence of these potential associations with sustained connectivity changes. Further, given that these associations with acutely altered conscious states were dose-adjusted, these subjective experiences may represent a correlate with sustained connectivity changes beyond what can be explained by dose alone.

Overall group-level analyses, without modelling these covariates, found fewer resting-state functional connectivity changes (with none surviving correction) relative to prior studies reporting either acute effects or sustained changes following higher psilocybin doses. For example, in a study of the acute effects of psilocybin 2 mg intravenously (equivalent to a low-strength oral dose) in fifteen healthy participants, three decreases and 30 increases in between-network functional connectivity were identified (Roseman et al., 2014). Three comparable neuroimaging studies explored sustained effects of oral psilocybin in healthy individuals, albeit at higher doses (25 mg/70 kg, n=12; 0.2-0.3 mg/kg, n=10; and 25 mg, n=7) (Barrett et al., 2020; McCulloch et al., 2022a; Siegel et al., 2024). This included increases in functional connectivity for 38 edges and decreases for 10 edges at one week and fewer changes still at one month (Barrett et al., 2020); decreased functional connectivity within the executive control network at one week and no changes at three months (McCulloch et al., 2022a), and decreased hippocampal connectivity with the default mode network (uncorrected) at three weeks which returned to baseline at 6-12 months (Siegel et al., 2024). Direct comparisons across resting-state fMRI studies are not straightforward, given disparate analysis methods (McCulloch et al., 2022b). Notwithstanding these caveats, our findings may suggest diminishing connectivity changes following lower doses as time passes.

This lack of significant changes observed in our study when aggregated at the group-level may, in part, reflect divergent directional effects across both dose and acutely altered conscious states, possibly cancelling each other out when combining across participants. All significant within- and between-network associations with these covariates were positive, such that lower doses and less intense alterations in conscious states were often associated with decreased connectivity at follow-up, whereas higher doses and greater alterations in conscious states were linked to increased connectivity. Replication in larger samples and clinical populations is required to substantiate these effects. If replicated, such directional patterns may inform design considerations in specific clinical cohorts, depending on the desired connectivity change.

This study’s strengths included randomization and blinding to the order of dosing, likely mitigating expectancy effects. This was evident as only 40% of participants correctly guessed the dose preceding their follow-up scan, which was comparable to chance performance (33%). Given the concerns for inflated effect sizes in psychedelic studies arising from placebo and expectancy confounds (Muthukumaraswamy et al., 2021), accompanied by known changes in brain activity (Romanella et al., 2023; Schienle and Wabnegger, 2024; Zilcha-Mano et al., 2019), the blinding efficacy improves the reliability of these neuroimaging findings.

The small sample size likely reduced the statistical power to identify subtle effects and rendered subgroup analyses at each dose not feasible. It also limits the generalisability of our findings, which should therefore be interpreted as exploratory and hypothesis-generating for future studies. While encouraging blinding efficacy was demonstrated, no placebo was offered. Therefore, a degree of expectancy related to receiving some dose of psilocybin was likely and could not be fully controlled for in this study.

## Conclusion

To our knowledge, this is the first study investigating the effects of varying dose levels and acutely altered conscious states on sustained, global changes in resting-state functional connectivity. Studies with larger sample sizes using similar dosing regimens are required to further investigate sustained changes, mediating factors, and corresponding treatment implications. If replicated, these positive associations may inform study design considerations in specific clinical cohorts.

## Acknowledgements

The authors wish to thank the Melbourne Brain Centre, Austin Health (Australia), for their support and assistance with MRI data acquisition.

## Authors’ contributions

RK is the Principal Investigator and the senior researcher. CB, OC, AB, and RK contributed to the development of the protocol. CB collected and analyzed the data. All authors contributed to the interpretation of the findings. CB drafted the manuscript. OD and RK provided supervision. All authors approved the final manuscript.

## Statements and Declarations

### Ethical considerations

Ethical approval for this study was granted by the Austin Health Human Research Ethics Committee (HREC/57390/Austin-2020). All procedures complied with the ethical standards of relevant national and institutional committees on human experimentation.

### Consent to participate

Informed consent was obtained from all participants for study participation.

### Consent for publication

Informed consent was obtained from all participants for the publication of their deidentified data.

## Funding statement

This work was supported by the Medical Research Future Fund (RK, OC, and AB; grant number MRF2012410) and the RANZCP Foundation, the Royal Australian and New Zealand College of Psychiatrists (CB).

The Usona Institute provided the study drug. They have not offered or provided payments to the investigators.

## Data availability

The data that support this study’s findings may be made available from the corresponding author upon reasonable request.

