## Supplementary Information for "Sustained Effects of Low-to-Moderate Doses of Psilocybin on Brain Connectivity"

**Neuroimaging Methods**

**Neuroimaging acquisition**

Images were obtained on a Siemens 3 Tesla wide-bore Vida MRI system fitted with a 64-channel BioMatrix head coil. Participants first underwent a localizer scan to allow the scanner to determine brain position within the magnet.

This was followed by a high-resolution magnetization-prepared rapid gradient echo (MP-RAGE) T1-weighted structural scan for functional time series co-registration. The acquisition parameters were: three-dimensional Fourier transform (3DFT) rapid spoiled gradient echo sequence; generalized autocalibrating partial parallel acquisition (GRAPPA) acceleration factor = 2; field of view (FOV) = 230.00 mm, matrix size = 256 x 256; slice thickness = 0.90 mm; slice gap = 0.00 mm; repetition time (TR) = 1970.00 ms; echo time (TE) = 2.12 ms; flip angle = 8 degrees; orientation = sagittal; phase encoding direction = anterior to posterior (A >> P); volume coverage in terms of Z = 172.80 mm; voxel size = 0.90 mm^3^; whole brain coverage; bandwidth = 220 Hz/Px; slices per slab = 192; order of slice acquisition: multi-slice mode = single shot, series = ascending; acquisition time = 4 minutes 43 seconds.

This was followed by two 60-second field maps, each differentiated by opposing phase encoding directions (A >> P, then P >> A) to correct distortions caused by magnetic field inhomogeneities in both the structural and resting-state fMRI scans.

Participants then completed a T2*-weighted resting-state functional MRI (fMRI) scan with their eyes closed and without music. The acquisition parameters were: single shot, two-dimensional echo-planar imaging sequence for blood oxygen level-dependent (BOLD) imaging sequence; GRAPPA acceleration factor = 2; FOV = 216.00 mm; matrix size = 72 x 72; slice thickness = 3.00 mm; slice gap = 0.00 mm; TR = 3000.00 ms; TE = 30.00 ms; flip angle = 85 degrees; orientation = 30.00 degrees oblique relative to the transverse plane (T > C-30.0); phase encoding direction = anterior to posterior (A >> P); volume coverage in terms of Z = 132.00 mm; voxel size = 3.00mm^3^; whole brain coverage; bandwidth = 2240 Hz/Px; number of slices = 44; order of slice acquisition: interleaved; acquisition time = 15 minutes.

To reduce head movement during scanning, headphones and foam pads were placed on the sides of the participants’ heads.

**Neuroimaging quality control and preprocessing**

Images from all participants were converted from the native Digital Imaging and Communications in Medicine (DICOM) format to the Neuroimaging Informatics Technology Initiative (NIfTI) standard file format and uploaded to the Distributed and Reflective Informatics System (DaRIS) <https://daris.researchsoftware.unimelb.edu.au>. Each subject’s data was visually inspected for obvious artefacts, signal drop-out or gross lesions.

Image preprocessing was performed in Statistical Parametric Mapping software (SPM12) running in Matlab (MathWorks, Natick, MA, USA). Preprocessing included head-motion correction, co-registration with the T1 structural weighted image, and then normalization to a common standard space defined by the Montreal Neurological Institute 152 template (Mazziotta et al., 2001). Skull stripping and segmentation of tissue types were undertaken for the anatomical images using the SPM intensity-based segmentation algorithm. The resultant combined white matter and cerebrospinal fluid (CSF) mask was subtracted from the gray matter mask with a stringent threshold of 1% so that voxels containing more than 1% white matter or CSF were removed. The resultant combined white matter and CSF mask was used to extract spurious signal from the data and was subjected to principal component analyses using a CompCor method (Behzadi et al., 2007). The first 5 components were retained from each analysis. A linear regression model that included these 10 component signals and the 6 head motion parameters (3 rotation, 3 translation) estimated during the head motion correction procedure, and the first-order derivatives of all 16 signals were fitted on a voxel-wise basis (Muschelli et al., 2014). Signal from white matter, cerebrospinal fluid, and the global brain signal, in addition to the 6 head motion parameters and their first derivatives, were modelled as nuisance variables and removed from the analysis. Regular visual quality control to ensure correct co-registration and normalization was conducted.

The functional data underwent linear detrending and bandpass filtering at 0.008-0.08 Hz to remove slow drifts in the BOLD signal often arising from fluctuations in physiological parameters such as heartbeat and respiration, scanner instabilities, and head motion. The data then underwent spatial smoothing with a Gaussian kernel (full width at half maximum) of 8 mm.

**Supplemental Table S1: Region of interest (ROI) labels**

| **ROI Number** | **Label [Network]** |
| --- | --- |
| 1 | LH Cingulate Cortex [Frontoparietal Control] |
| 2 | LH Orbitofrontal Cortex [Frontoparietal Control] |
| 3 | LH Dorsal Prefrontal Cortex [Frontoparietal Control] |
| 4 | LH Lateral Prefrontal Cortex [Frontoparietal Control] |
| 5 | LH Medial Prefrontal Cortex (Posterior) [Frontoparietal Control] |
| 6 | LH Ventral Prefrontal Cortex [Frontoparietal Control] |
| 7 | LH Parietal Cortex [Frontoparietal Control] |
| 8 | LH Temporal Cortex [Frontoparietal Control] |
| 9 | LH Precuneus [Frontoparietal Control] |
| 10 | RH Cingulate Cortex [Frontoparietal Control] |
| 11 | RH Lateral Prefrontal Cortex [Frontoparietal Control] |
| 12 | RH Medial Prefrontal Cortex (Posterior) [Frontoparietal Control] |
| 13 | RH Ventral Prefrontal Cortex [Frontoparietal Control] |
| 14 | RH Parietal Cortex [Frontoparietal Control] |
| 15 | RH Temporal Cortex [Frontoparietal Control] |
| 16 | RH Precuneus [Frontoparietal Control] |
| 17 | LH Prefrontal Cortex [Default Mode] |
| 18 | LH Parahippocampal Cortex [Default Mode] |
| 19 | LH Parietal Cortex [Default Mode] |
| 20 | LH Temporal Cortex [Default Mode] |
| 21 | LH Precuneus / Posterior Cingulate Cortex [Default Mode] |
| 22 | RH Dorsal/Medial Prefrontal Cortex [Default Mode] |
| 23 | RH Ventral Prefrontal Cortex [Default Mode] |
| 24 | RH Parietal Cortex [Default Mode] |
| 25 | RH Temporal Cortex [Default Mode] |
| 26 | RH Precuneus / Posterior Cingulate Cortex [Default Mode] |
| 27 | LH Frontal Eye Fields [Dorsal Attention] |
| 28 | LH Posterior Parietal Cortex [Dorsal Attention] |
| 29 | LH Precentral Cortex [Dorsal Attention] |
| 30 | RH Frontal Eye Fields [Dorsal Attention] |
| 31 | RH Posterior Parietal Cortex [Dorsal Attention] |
| 32 | RH Precentral Cortex [Dorsal Attention] |
| 33 | LH Frontal Operculum / Insula [Salience] |
| 34 | LH Medial Cortex [Salience] |
| 35 | LH Lateral Prefrontal Cortex [Salience] |
| 36 | LH Parietal Operculum [Salience] |
| 37 | LH Temporo-Occipital Cortex [Salience] |
| 38 | RH Frontal Operculum / Insula [Salience] |
| 39 | RH Medial Cortex [Salience] |
| 40 | RH Lateral Prefrontal Cortex [Salience] |
| 41 | RH Ventral Prefrontal Cortex [Salience] |
| 42 | RH Precentral Cortex [Salience] |
| 43 | RH Temporo-Occipital / Parietal Cortex [Salience] |
| 44 | LH Somatomotor Region [Somatomotor] |
| 45 | RH Somatomotor Region [Somatomotor] |
| 46 | LH Visual Cortex [Visual] |
| 47 | RH Visual Cortex [Visual] |
| 48 | LH Orbitofrontal Cortex [Limbic] |
| 49 | LH Temporal Pole [Limbic] |
| 50 | RH Orbitofrontal Cortex [Limbic] |
| 51 | RH Temporal Pole [Limbic] |

LH = Left hemisphere

RH = Right hemisphere

**Results**

**Supplemental Table S2: Participants**

| **A: Demographics** (Total, n = 10) | |
| --- | --- |
| **Age** [Mean (range)] (years) | 30.1 (22-39) |
| **Height** [Mean (range)] (cm) | 170.8 (160-188) |
| **Weight** [Mean (range)] (kg) | 68.5 (51-94) |
| **Gender** |  |
| Male | 5 (50%) |
| Female | 5 (50%) |

Data are n (%) unless otherwise specified.

| **B: Dose and resting-state fMRI timepoint** | | | |
| --- | --- | --- | --- |
| **Participant** | **Dose before follow-up scan (mg)** | **Number of doses before follow-up scan** | **Days between dose and follow-up scan** |
| 1 | 10 | 1 | 7 |
| 2 | 5 | 1 | 7 |
| 3 | 10 | 1 | 7 |
| 4 | 20 | 2 | 7 |
| 5 | 15 | 2 | 7 |
| 6 | 20 | 2 | 6 |
| 7 | 20 | 1 | 7 |
| 8 | 20 | 1 | 7 |
| 9 | 15 | 1 | 7 |
| 10 | 10 | 1 | 7 |

**Supplemental Figure S1: CONSORT flowchart**

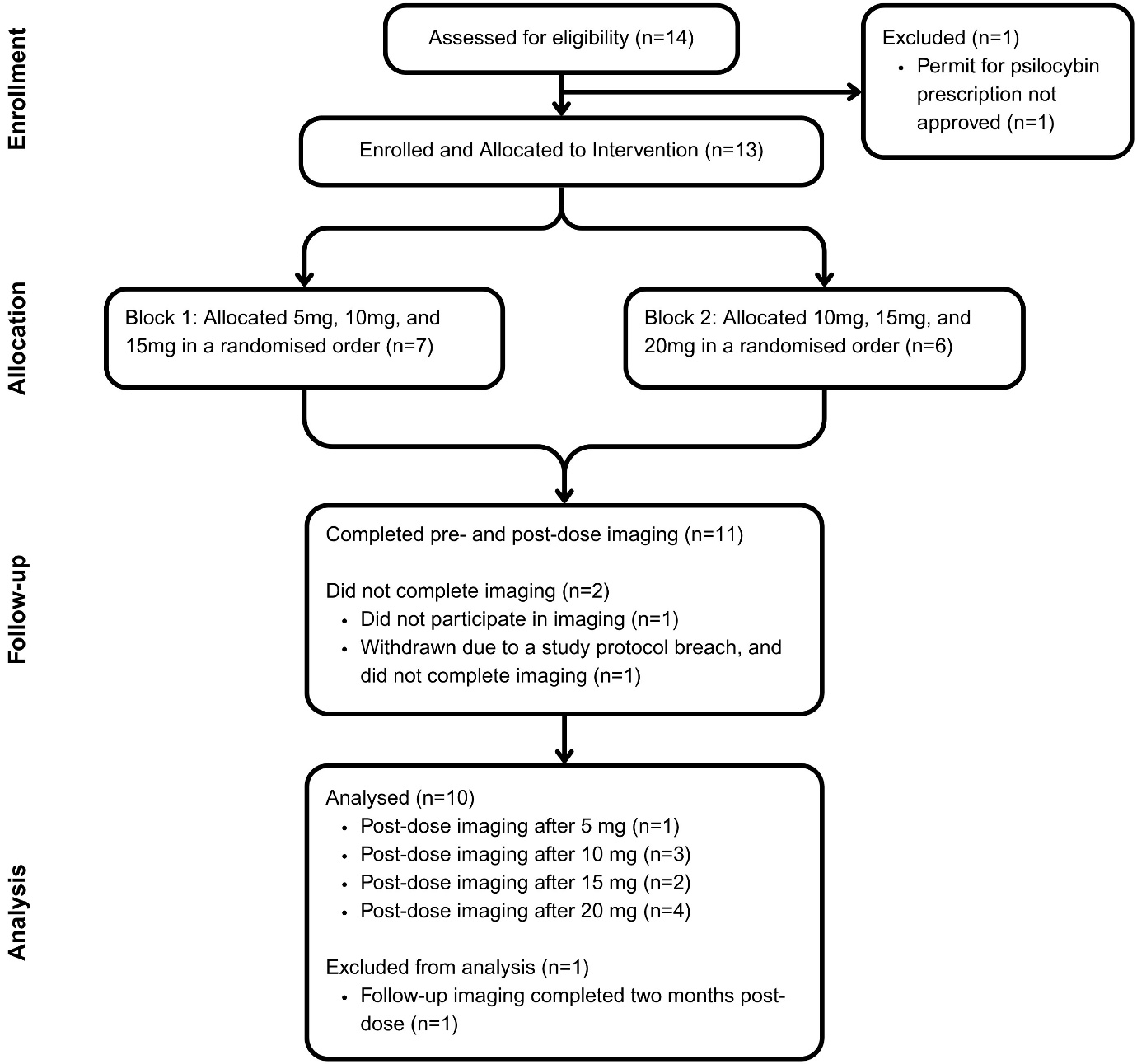

**Supplemental Figure S2: Associations (dose-adjusted) between 5-Dimensional Altered States of Consciousness, Ego-Dissolution Inventory, and edgewise functional connectivity change**

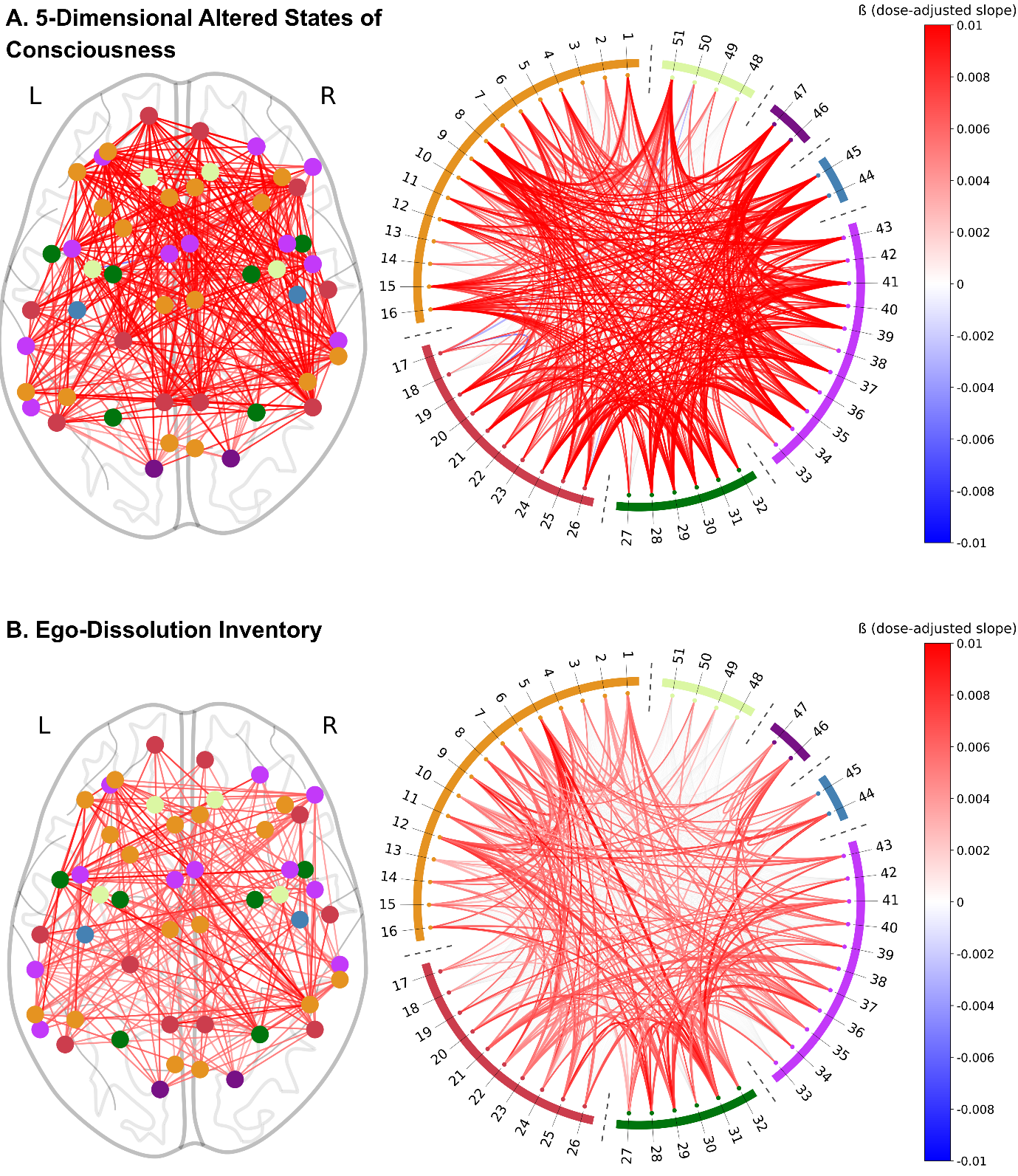

Edgewise functional connectivity changes at follow-up compared to baseline as a function of 5-Dimensional Altered States of Consciousness (A) and mean Ego-Dissolution Inventory (B) (dose-adjusted). Results are viewed at *p_unc_* < 0.05. Region of interest (ROI) color is represented by its corresponding network. Edge color is represented by the dose-response slope (β) per 5mg (see color bar on the right). Relative edge width is represented by the *t*-statistic. See Table S1 for ROI labels.

**Supplemental Figure S3: Associations (unadjusted) between 5-Dimensional Altered States of Consciousness, Ego-Dissolution Inventory, and network-level functional connectivity change**

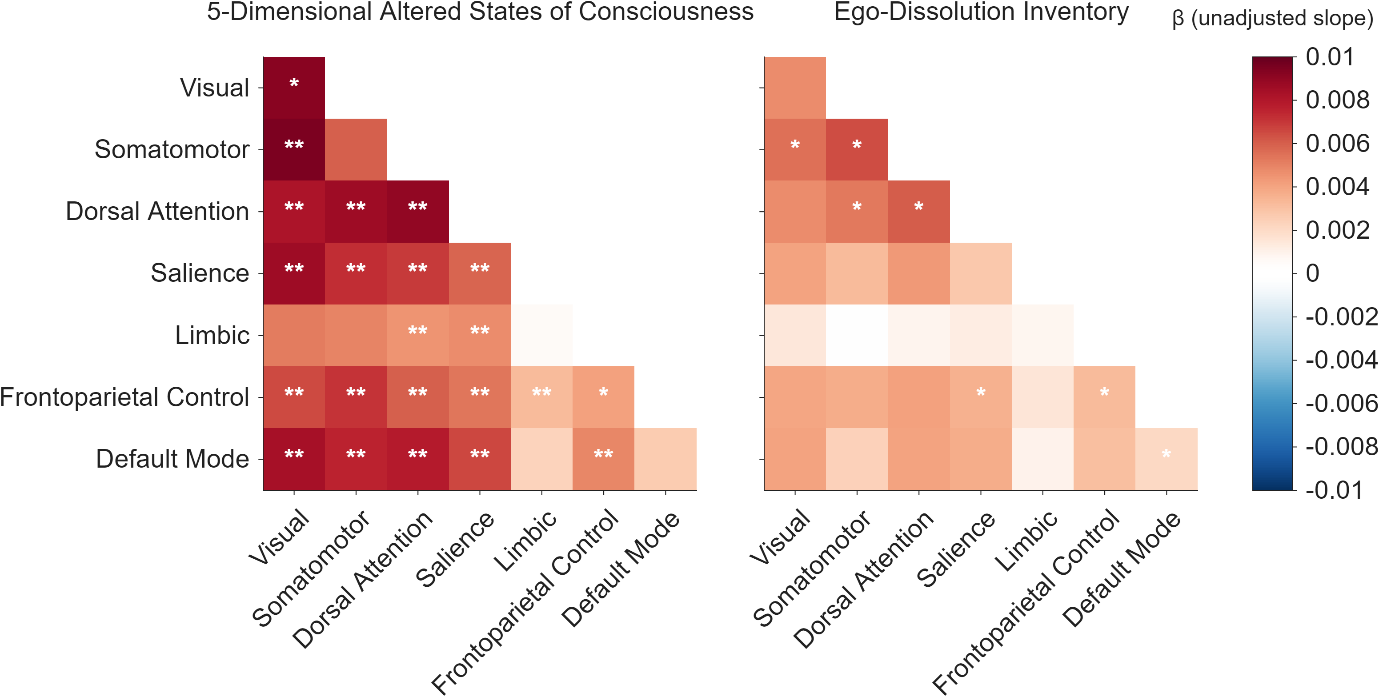

Square color is represented by the slope (β) (see color bar on the right). The diagonal squares represent within-network change, while the off-diagonal squares represent between-network change. **p*_unc_ < 0.05; ***p*_FDR_ < 0.05.

**Supplemental Figure S4: Associations between dose and network-level functional connectivity change**

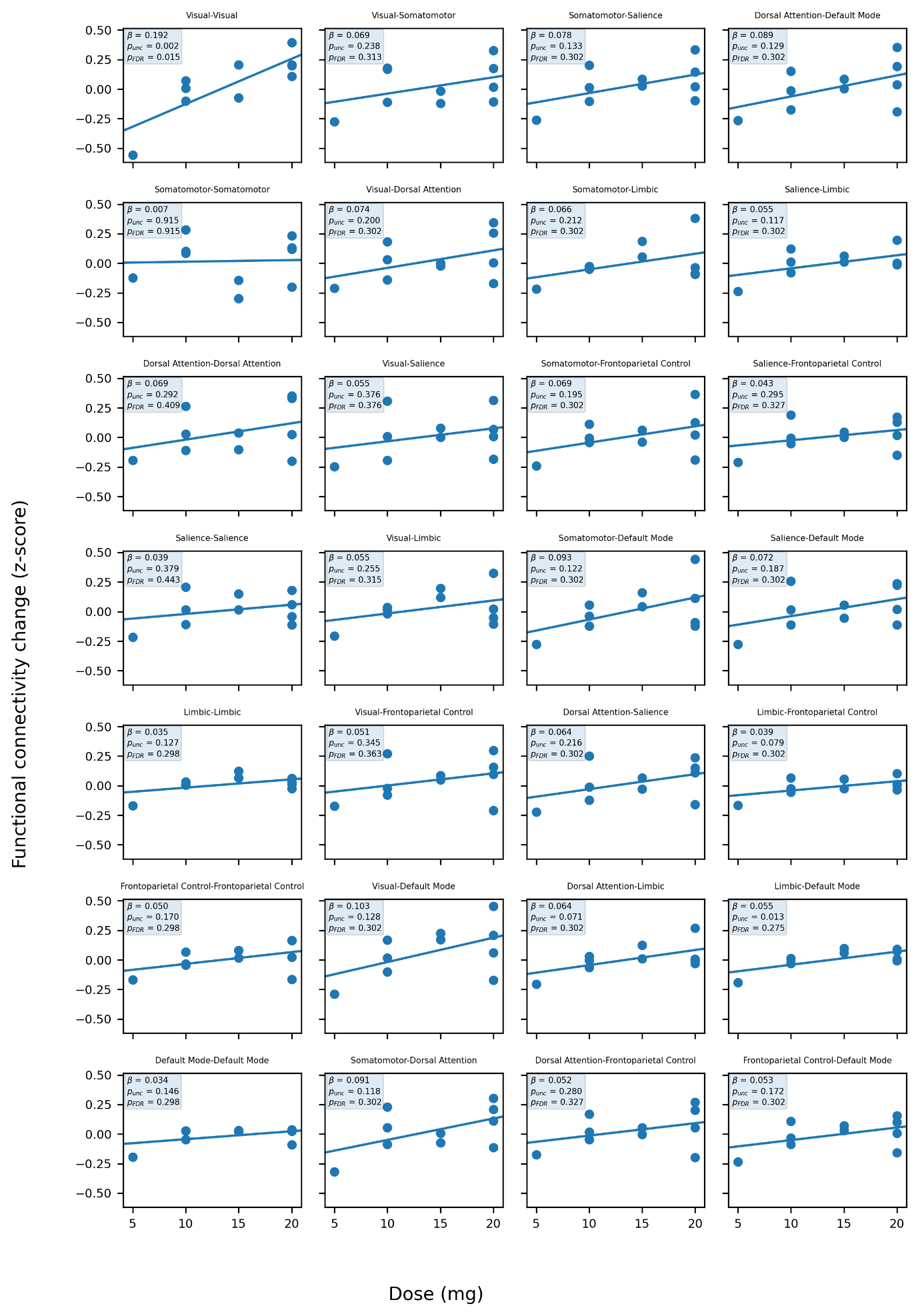

**2**

**Supplemental Figure S5: Associations between 5-Dimensional Altered States of Consciousness and network-level functional connectivity change (dose-adjusted)**

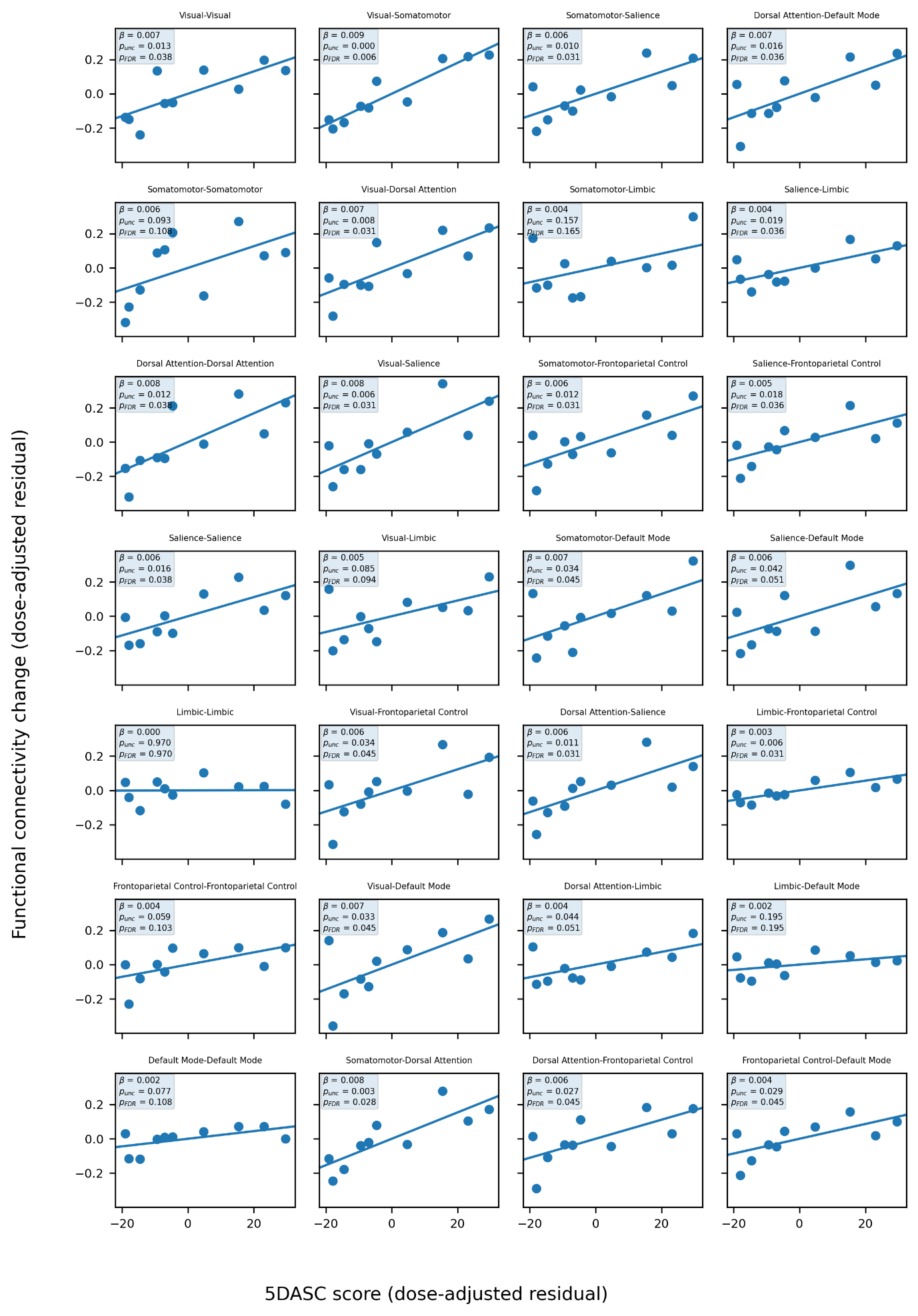

5DASC = 5-Dimensional Altered States of Consciousness

**Supplemental Figure S6: Associations between Ego-Dissolution Inventory (EDI) and network-level functional connectivity change (dose-adjusted)**

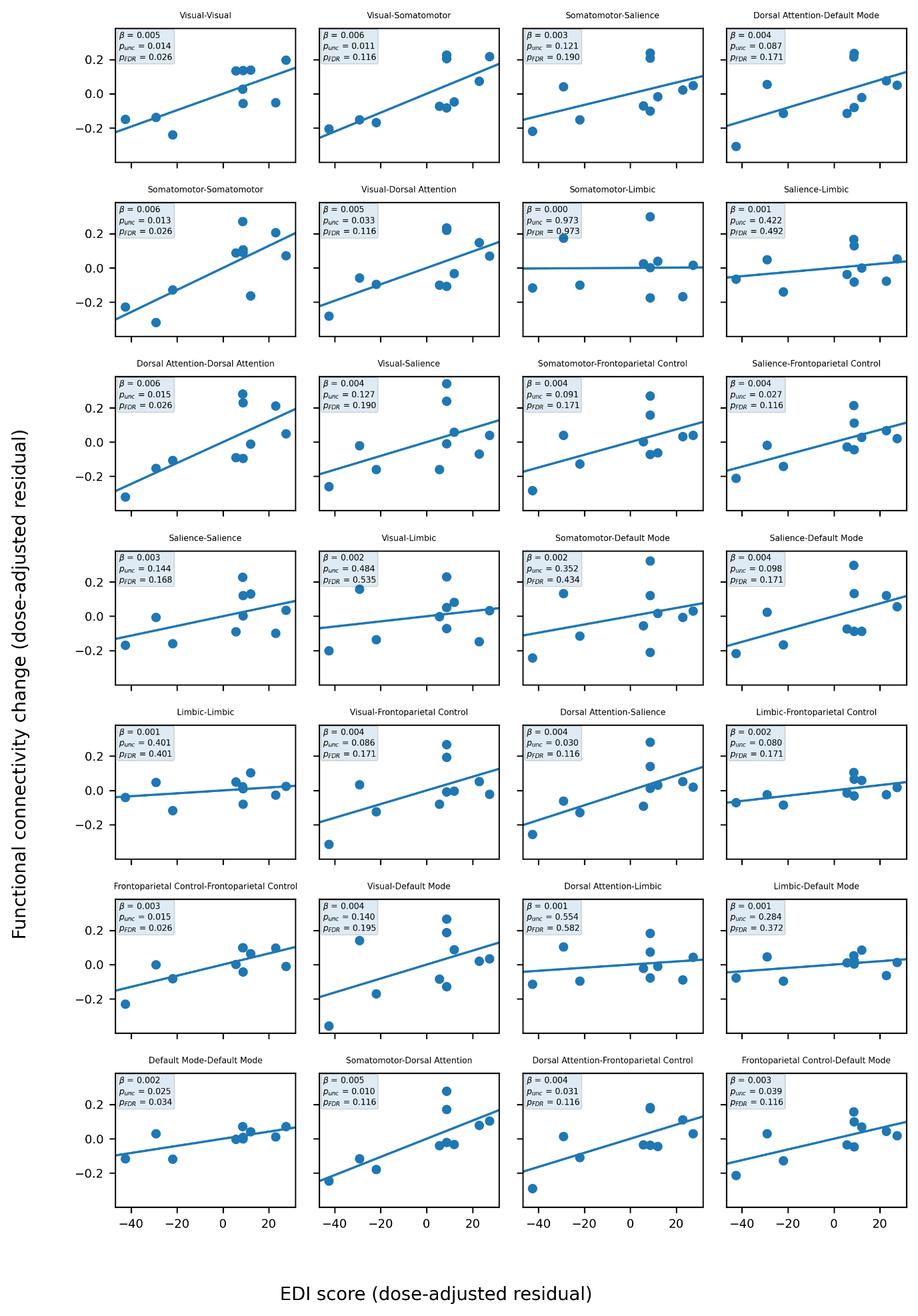

EDI = Ego-Dissolution Inventory

**Supplemental Figure S7: Group-level functional connectivity changes**

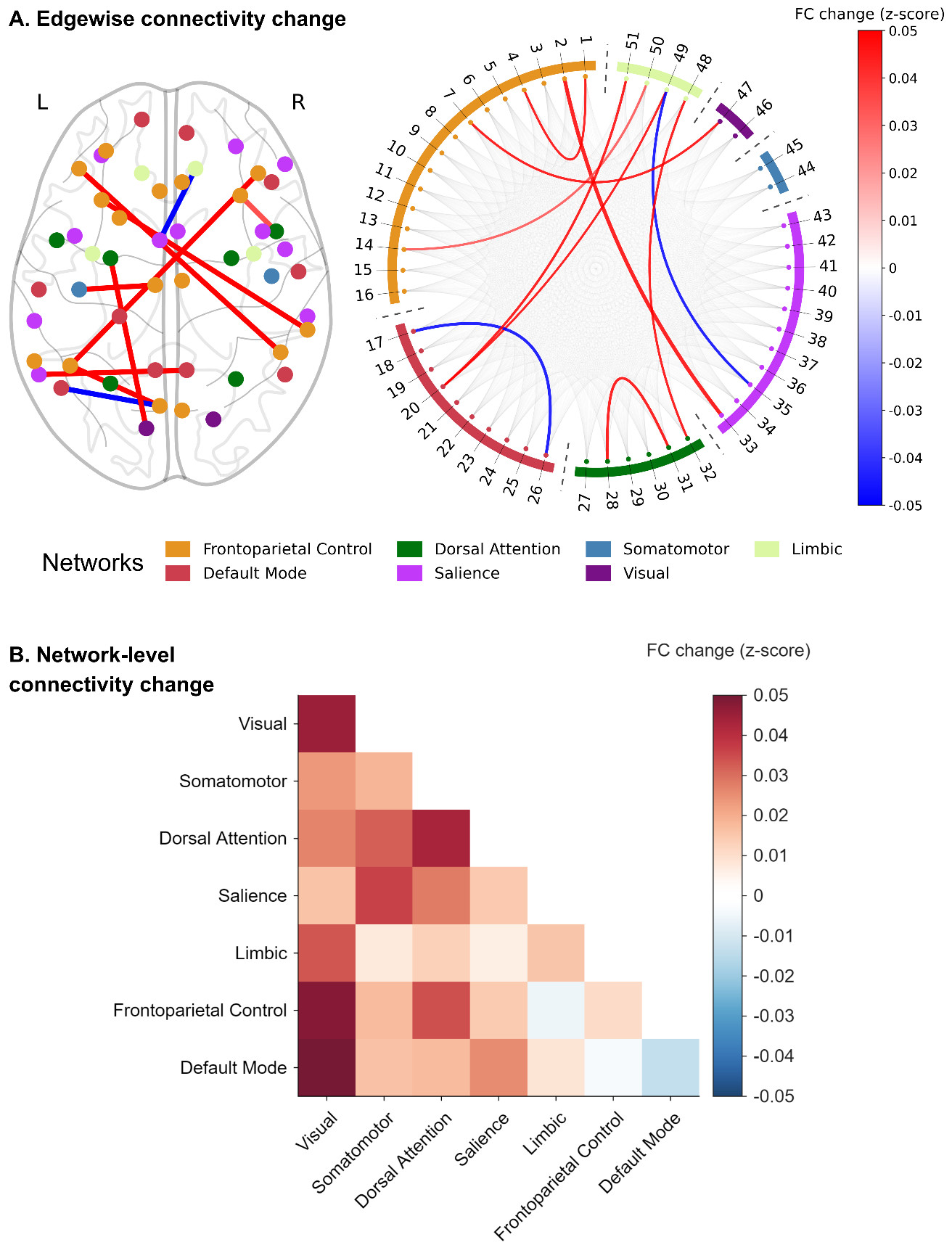

A: Edgewise functional connectivity changes at follow-up compared to baseline. Results are viewed at *p_unc_* < 0.05. Region of interest (ROI) color is represented by its corresponding network. Edge color is represented by the mean functional connectivity change (FC change (z-score)) (see color bar on the right). Relative edge width is represented by the *t*-statistic. See Table S1 for ROI labels.

B: Network-level functional connectivity changes at follow-up compared to baseline. Square color is represented by the mean FC change (z-score) (see color bar on the right). The diagonal squares represent within-network change, while the off-diagonal squares represent between-network change. No squares survived *p_unc_* < 0.05.

FC = Functional connectivity
